# U.S. adolescents’ mental health and COVID-19-related changes in technology use, Fall 2020

**DOI:** 10.1101/2021.03.15.21253598

**Authors:** Taylor A. Burke, Emily R. Kutok, Shira Dunsiger, Nicole R. Nugent, John V. Patena, Alison Riese, Megan L. Ranney

## Abstract

Preliminary reports suggest that during COVID-19, adolescents’ mental health has worsened while technology and social media use has increased. Much data derives from early in the pandemic, when schools were uniformly remote and personal/family stressors related to the pandemic were limited. This cross-sectional study, conducted during Fall 2020, examines the correlation between mental wellbeing and COVID-19-related changes in technology use, along with influence of COVID-19-related stressors, school status (in-person versus remote), and social media use for coping purposes, among 978 U.S. adolescents. Results suggest self-reported daily social media and technology use increased significantly from prior to COVID-19 through Fall 2020. Increased social media use was significantly associated with higher levels of anxiety and depressive symptoms regardless of other theoretical moderators or confounders of mental health (e.g., demographics, school status, importance of technology, COVID-19-related stress). Despite literature suggesting that remote learning may result in adverse mental health outcomes, we did not find local school reopening to be associated with current depressive/anxiety symptoms, nor with COVID-19-related increases in technology use. Self-reported use of social media for coping purposes moderated the association between increased social media use and mental health symptoms; in other words, some social media use may have positive effects. Although much prior research has focused on social media use as a marker of stress, we also found that increased video gaming and TV/movie watching were also associated with internalizing symptoms, in accordance with others' work. Future research should explore in more granular detail what, if any, social media and technology use is protective during a pandemic, and for whom, to help tailor prevention efforts.

## INTRODUCTION

Preliminary reports suggest that during COVID-19, adolescents’ mental health has worsened while technology and social media use has increased. Much data derives from early in the pandemic, when schools were uniformly remote and personal/family stressors related to the pandemic were limited. This cross-sectional study, conducted during Fall 2020, examines the correlation between mental wellbeing and COVID-19-related changes in technology use, along with influence of COVID-19-related stressors, school status (in-person versus remote), and social media use for coping purposes, among U.S. adolescents.

## METHODS

From September 23 to December 16, 2020, English-speaking adolescents (ages 13-17) residing in the United States were recruited using Instagram for an online survey, with approval from the Institutional Review Board. Assent was waived, with approval from the Institutional Review Board. Self-report measures (adapted from Pew Internet Survey^1^) assessed average daily duration of technology use (social media, phone/video calls, video games, TV/movie/videos) 30 days before initial COVID-19-related school closures versus past week. Standard measures for past week anxiety and depressive symptoms (PROMIS)^2^, well-being (WHO-5)^3^, and cybervictimization^4^ were used. Use of social media for coping through social connection was assessed using an adapted measure for the purpose of the present study. School status (open full-time or hybrid versus closed) was determined through the use of the COVID-19 US State Policy Database.^5^ COVID-19-related stressors^6^, perceived importance of social media^7^, and demographics were also assessed.

Generalized linear models were used to examine associations between changes in technology use and current mental health outcomes, adjusting for COVID-19-related stressors and importance of social media (identified as confounders in preliminary analysis); potential moderators were examined.

## RESULTS

We recruited 978 youth from 42 states (Table 1). All forms of technology use significantly increased from pre-COVID until the time of assessment (Table 2a). After adjustment for confounders, self-reported increases in social media use were associated with higher anxiety (b = .07, SE = .03, *p* = .02) and depressive symptoms (b = .11, SE = .03, *p* < .001) (Table 2b). Low use of social media for coping moderated the association between social media use and depressive symptoms (b = .15, SE = 0.07, *p* = .02). Increases in video gaming and TV/movie watching were associated with higher depressive symptoms (b = .06, SE = .03, *p* = .04; b = .10, SE = .03, *p* = .002), and video gaming was associated with higher anxiety (b = .09, SE = .03, *p* = .01).

**Table 1.** Participant demographics and clinical characteristics (N = 978)

| Characteristic | Mean (SD) or n (%) |
| --- | --- |
| Age | 15.16 (1.31) |
| Gender |  |
| Female | 568 (58.1) |
| Male | 197 (20.1) |
| Transgender | 46 (4.7) |
| Not Listed | 135 (13.8) |
| Prefer not to answer | 32 (3.3) |
| Sexual orientation |  |
| Straight | 328 (33.5) |
| Gay or lesbian | 128 (13.1) |
| Bisexual | 321 (32.8) |
| Other | 156 (16.0) |
| Prefer not to Answer | 45 (4.6) |
| Race |  |
| American Indian/Alaska Native | 6 (0.6) |
| Asian | 73 (7.5) |
| Native Hawaiian or other<br>pacific islander | 2 (0.2) |
| Black or African American | 36 (3.7) |
| White | 712 (72.8) |
| Multiple categories | 97 (9.8) |
| Other | 29 (3.0) |
| Prefer not to answer | 23 (2.4) |
| Ethnicity |  |
| Hispanic or Latino |  |
| Yes | 105 (10.7) |
| No | 851 (87.0) |
| Unknown | 16 (1.6) |
| Prefer not to answer | 6 (0.6) |
| SES: Low | 336 (34.4) |
| Region |  |
| NE | 68 (7.0) |
| NW | 67 (6.9) |
| SW | 140 (14.3) |
| SE | 209 (21.4) |
| Midwest | 191 (19.5) |
| West | 100 (10.2) |
| Valid Zipcode not provided | 203 (20.8) |
| Open schools (full-time or hybrid) at the time of study completion* | 558 (72.1) |
| Own smartphone | 936 (98.4) |
| Number of COVID stressors | 7.36 (3.75) |

\*School status was assessed via COVID-19 US State Policy Database<sup>7</sup> that tracks the dates when each US state implemented new social safety net, economic, and physical distancing policies in response to the COVID-19 pandemic. This database was developed and maintained by researchers at the Boston University School of Public Health, and is updated at least biweekly. We determined public school reopening in the participant's reported zipcode for the two weeks prior to survey completion.

**Table 2a.**
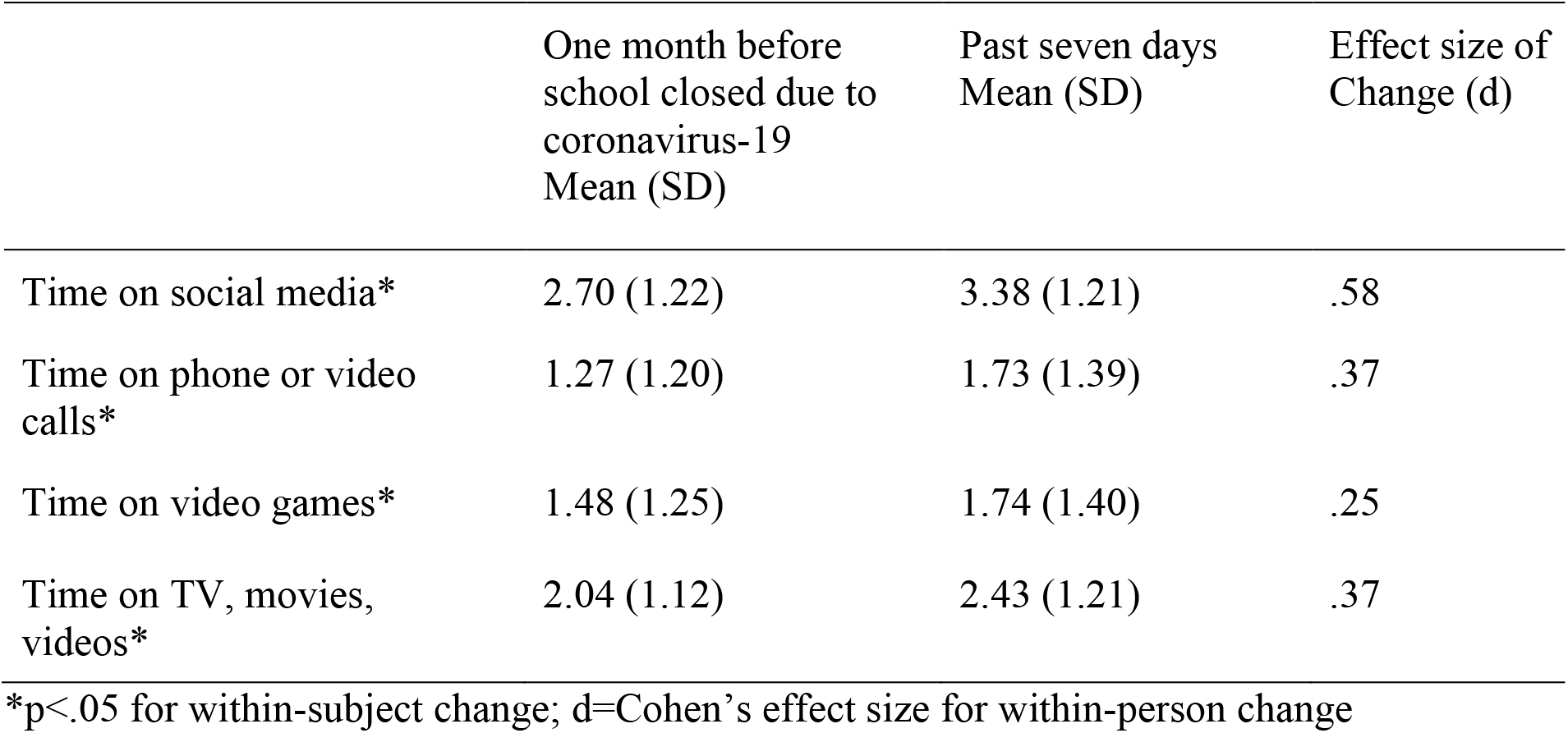
Within-person self-reported daily technology use (in hours), one month prior to school closures, compared to the 7 days prior to the survey

|  | One month before<br>school closed due to<br>coronavirus-19<br>Mean (SD) | Past seven days<br>Mean (SD) | Effect size of<br>Change (d) |
| --- | --- | --- | --- |
| Time on social media* | 2.70 (1.22) | 3.38 (1.21) | .58 |
| Time on phone or video<br>calls* | 1.27 (1.20) | 1.73 (1.39) | .37 |
| Time on video games* | 1.48 (1.25) | 1.74 (1.40) | .25 |
| Time on TV, movies,<br>videos* | 2.04 (1.12) | 2.43 (1.21) | .37 |

\*p<.05 for within-subject change; d=Cohen's effect size for within-person change

**Table 2b.** Adjusted effects of changes in technology use on mental health outcomes

|  | Anxiety<br>Symptoms | Depressive<br>Symptoms | Well-being | Cybervictimization |
| --- | --- | --- | --- | --- |
| Change in time on social<br>media | b=.07, SE=.03* | b=.11, SE=.03* | b=-.21, SE=.13 | b=-.06, SE=.08 |
| Change in time on<br>phone or video calls | b=.02, SE=.03 | b=.02, SE=.03 | b=.07, SE=.12 | b=-.08, SE=.07 |
| Change in time on video<br>games | b=.09, SE=.03* | b=.06, SE=.03* | b=-.03, SE=.14 | b=.12, SE=.09 |
| Change in time on TV,<br>movies, videos | b=.06, SE=.03 | b=.10, SE=.03* | b=-.05, SE=.15 | b=-.06, SE=.09 |

Note. Changes in technology time reflect differences: past 7 days – one month before school closures; Models adjusted for COVID-19-specific stressors and importance of social media; b=unstandardized regression coefficient; SE=standard error. \*p<.05

There were no associations between changes in any form of technology use and well-being or cybervictimization. Neither local school status, nor level of COVID-19-related stressors, nor self-perceived importance of technology, were significant confounders or moderators of the observed effect.

## DISCUSSION

In this geographically diverse sample of adolescents across the United States, self-reported daily social media and technology use increased significantly from prior to COVID-19 through Fall 2020. Increased social media use was significantly associated with higher levels of anxiety and depressive symptoms regardless of other theoretical moderators or confounders of mental health (e.g., demographics, school status, importance of technology, COVID-19-related stress). Despite literature suggesting that remote learning may result in adverse mental health outcomes^8^, we did not find local school reopening to be associated with current depressive/anxiety symptoms, nor with COVID-19-related increases in technology use. Self-reported use of social media for coping purposes moderated the association between increased social media use and mental health symptoms; in other words, some social media use may have positive effects.^9^ Although much prior research has focused on social media use as a marker of stress, we also found that increased video gaming and TV/movie watching were also associated with internalizing symptoms, in accordance with others’ work.^10^ Future research should explore in more granular detail what, if any, social media and technology use is protective during a pandemic, and for whom, to help tailor prevention efforts. Limitations include use of some non-validated measures, reliance on self-report of technology use, and use of a national database to assess school status.

In conclusion, our study shows that, although adolescents’ self-reported technology use increased from prior to the pandemic until Fall 2020 and was associated with poorer mental health, the relationship may be more nuanced than previously reported.

## Data Availability

Data may be made available upon request.

## Acknowledgments

The content is solely the responsibility of the authors and does not necessarily represent the official views of the university or the TAM program. Taylor Burke supported by NIMH T32 MH019927.

